# Detection of COVID-19 Disease from Chest X-Ray Images: A Deep Transfer Learning Framework

**DOI:** 10.1101/2020.11.08.20227819

**Authors:** Shadman Sakib, Md. Abu Bakr Siddique, Mohammad Mahmudur Rahman Khan, Nowrin Yasmin, Anas Aziz, Madiha Chowdhury, Ihtyaz Kader Tasawar

## Abstract

World economy as well as public health have been facing a devastating effect caused by the disease termed as Coronavirus (COVID-19). A significant step of COVID-19 affected patient’s treatment is the faster and accurate detection of the disease which is the motivation of this study. In this paper, implementation of a deep transfer learning-based framework using a pre-trained network (ResNet-50) for detecting COVID-19 from the chest X-rays was done. Our dataset consists of 2905 chest X-ray images of three categories: COVID-19 affected (219 cases), Viral Pneumonia affected (1345 cases), and Normal Chest X-rays (1341 cases). The implemented neural network demonstrates significant performance in classifying the cases with an overall accuracy of 96%. Most importantly, the model has shown a significantly good performance over the current research-based methods in detecting the COVID-19 cases in the test dataset (Precision = 1.00, Recall = 1.00, F1-score = 1.00 and Specificity = 1.00). Therefore, our proposed approach can be adapted as a reliable method for faster and accurate COVID-19 affected case detection.

## I. Introduction

COVID-19, the pandemic that has brought the world to a halt, was reported in Wuhan, China in the December of 2019 for the first time, when patients with cases of unidentified pneumonia emerged. The virus responsible for the disease, named SARS-CoV-2, belongs to a family of coronaviruses that are zoonotic in nature. Until SARS-CoV-2 surfaced, six types of coronaviruses were known to be able to harm humans by mainly targeting the respiratory system. Among them, two had caused epidemics in the last two decades named SARS-CoV and MERS-CoV. Despite the mortality rates of these epidemics being much higher than that of COVID-19 (10% for SARS and 30-35% for MERS), the cumulative number of deaths for the latter has surpassed that of both the epidemics combined by many folds [1]. As of 28 June 2020, the total number of global cases and deaths exceed 9.8 million and 4.9 lakh respectively [2].

The common clinical characteristics of COVID-19 include a range of symptoms mutual with other viral diseases such as the common cold. In more severe or progressed cases, pneumonia, development of fluid in the lungs, acute respiratory distress syndrome (ARDS), multiorgan failure, septic shock as well as death may occur. Elderly people or people exhibiting comorbidity having a compromised immune system are highly prone to infection and severity. On the other hand, many individuals do not show any symptoms despite being carriers of the virus. This makes detection and containment of the virus even harder. Along with being a highly contagious disease, COVID-19 has a long incubation period, on average, five to six days between the contact and symptom onset phases. Thus, abiding by preventive measures such as social distancing, hygiene maintenance, and contact tracing and enabling a system that can diagnose the disease earlier and faster is paramount.

At present, the eminent standard for diagnosing COVID-19 is the reverse transcription-polymerase chain reaction (RT-PCR) which identifies the nucleotides of the virus from specimens extracted from a nasal swab or oropharyngeal swab. One of the major drawbacks of this method is the tedium involved and the time required as the fastest turn-around time is at least 24 hours. Added with the rapid spread and hence an increased number of specimens collected, the laboratories very rapidly get overwhelmed. Furthermore, it is laborious, relatively expensive, and has a low sensitivity (60%–70%) [3]. Many countries suffer from false results due to multiple plausible causes such as specimen handling, stage of disease when the specimen is collected, and quality of the specimen [4]. With limited resources i.e. testing kits, hospital beds and ICU beds, ventilators, personal protective equipment (PPE), the healthcare systems around most of the globe are loaded at the havoc and bound to make selective decisions in terms of testing, patient admission, ICU beds as well as the provision of ventilators.

Radiography chest images (X-ray and CT scan) analysis is a valuable alternative of the PCR method. They may assist in multiple ways from diagnosing the disease to sorting out the high-risk patients for quarantining and prioritizing while selective testing to identifying the false-negative PCR cases. However, since most viral cases of pneumonia’ images are akin and overlap, it is very difficult and time consuming for radiologists to distinguish the fine details by vision. Artificial Intelligence models can be a prompt solution. Very recently, the deep learning (DL) approach have been very popularly and successfully used in medical image classification applications owing to its powerful accuracy.

Some of the very recent works in detecting COVID-19 involves the application of various DL approaches. Due to the problem of COVID-19 being very recent, the unavailability of immense datasets has caused most of the works to use the technique called transfer learning. For instance, Ozturk *et al*. [5] presented a new model, DarkCovidNet which was a modified form of the Darknet-19 model for automatically detecting COVID-19 from raw images (Chest X-ray). 98.08% accuracy was achieved by their model for binary classification for indicating the presence of COVID-19 and 87.02% accuracy was obtained for multi-class classification between COVID-19 and pneumonia. In another study [6], three pre-trained deep CNN based models (Inception-ResNetV2, ResNet-50, and InceptionV3) were used for the automatic identification of COVID-19 pneumonia infected patients from the X-ray radiographs of the chest. Their results indicated that ResNet-50 displayed superior performance compared to the other two models. ResNet-50 showed an accuracy of 98%, InceptionV3 had 97% accuracy and 87% accuracy was shown by the other model. Hemdan *et al*. [7] introduced COVIDX-Net, a new DL framework that was created on the basis of seven different architectures of deep CNN (Xception, VGG19, MobileNetV2, DenseNet201, InceptionResNetV2, ResNetV2, and InceptionV3). Identifying COVID-19 from X-ray images (chest) by using this framework was the main objective. The best performance in this framework was displayed by DenseNet201 and VGG19 obtaining an f1 score of 0.91 for classifying COVID-19. Moreover, in a study [8] of detecting COVID-19 pneumonia as well as viral pneumonia, the researchers developed a public database where COVID-19 and viral pneumonia along with normal X-ray images of the chest were the constituents. Their work was to test four pre-trained CNNs namely, ResNet18, AlexNet, DenseNet201, and SqueezeNet on two schemes. One scheme was COVID-19 pneumonia and normal classification whereas the other scheme was COVID-19 pneumonia, viral pneumonia as well as normal classification. From SqueezNet, both the schemes achieved 98.3% accuracy which was the highest among the networks. Image augmentation played a vital role in obtaining such a level of accuracy. Farooq *et al*. [9] presented a work where an existing pre-trained architecture named ResNet-50 was fine-tuned for its performance improvement to identify COVID-19 and other pneumonia cases (bacterial, viral). The fine-tuned version of the architecture was called COVID-ResNet. 96.3% accuracy was achieved by their model. Furthermore, a study [10] to classify the images (chest X-ray) to viral pneumonia along with COVID-19 and normal cases was done. Their classifier was CheXNet based and transfer learning was implemented. Results indicated an accuracy of 97.8% of their presented model.

In this research, a deep convolutional neural network (DCNN) based on a pre-trained model for the automatic detection of COVID-19 from two other classes (Viral Pneumonia and normal chest X-ray images) was proposed. For this purpose, we used a fine-tuned ResNet-50 previously trained on the ImageNet dataset in our model. For the experiment, we used chest X-ray images rather than CT scans to fine-tuned the ResNet-50 model for classification. X-rays are relatively cheaper, quicker, lower patient dose, and more widely available in contrast to the expensive, higher radiation exposure and time-consuming CT scan machines and scans. Furthermore, portable X-ray machines allow for testing within an isolation ward, thus reducing risks of nosocomial infections and the number of PPEs used.

Organization of the remaining parts of the paper is as follows: Section II comprises of materials and methodology that provide details of the dataset containing X-ray images (chest), data pre-processing, details of dataset splitting, and the description of the proposed deep transfer learning network architecture. Experimental results and discussion, performance evaluation, and the result comparison with the current research-based methods are presented in Section III, and lastly, the conclusion of the paper resides in section IV.

## II. Materials and Methodology

Generally, DCNNs perform better in a greater database than a database which is comparatively smaller. Transfer learning is useful in such frameworks of CNN which have a relatively limited collection of data. In this study, we have undertaken the task of classifying images of chest X-rays into one of three classes: COVID-19 positive, viral pneumonia, or normal.

In our experiment, the collected database of X-ray images (chest) are initially made to undergo pre-processing and augmentation to increment the variation and make it more suitable for classification. The classification model is based on a version of a powerful pre-trained network used in tasks of computer vision, ResNet-50 [11]. ResNet-50, short for Residual Networks-50 is a 50-layer deep convolutional neural network. The concept of transfer learning is utilized in the model, where the weights of ResNet-50 trained on ImageNet is loaded and used for our task. ImageNet is a database spanned across 20,000+ categories of over 14 million images. The schematic representation of the proposed method’s overall workflow is depicted in Figure 1.

**Fig. 1.**
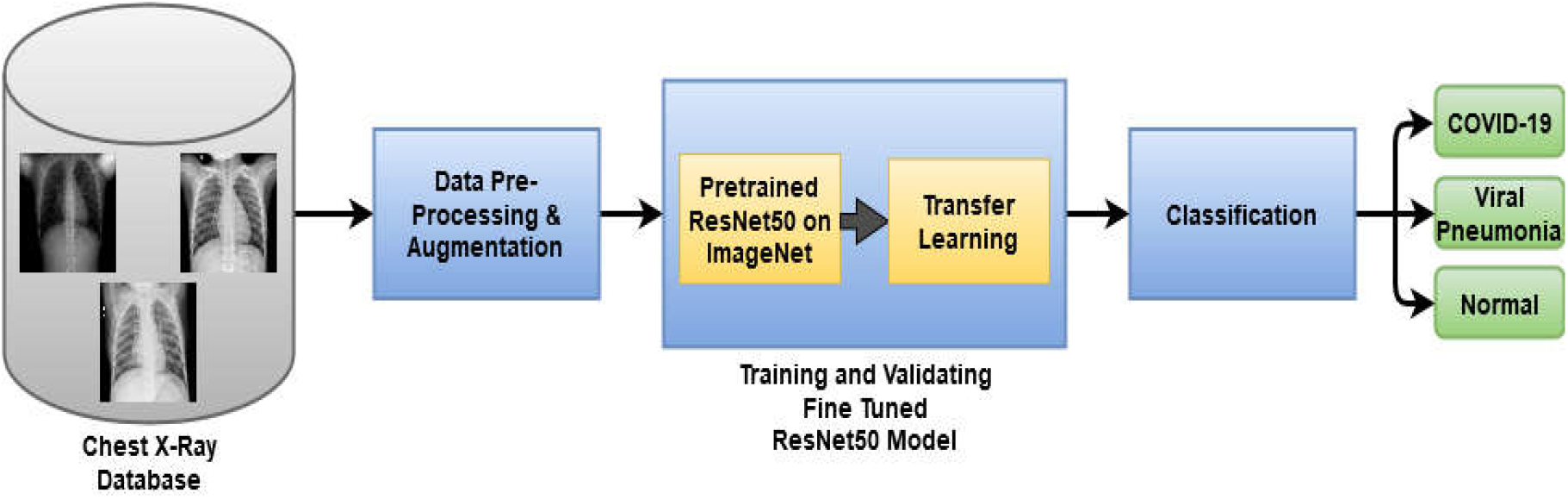
Workflow of proposed system for COVID-19 detection from chest X-ray images using Pre-trained ResNet50 and Transfer Learning

### A. Chest X-ray Database

The database used in this study consists of 2905 posterior-anterior (PA) or anterior-posterior (AP) images (chest X-ray) and is curated by Chowdhury et al. [8]. Each sample of the database is in Portable Network Graphics (PNG) format and is sized 1024×1024 pixels. They can be easily converted to conventional sizes of 224×224 or 227×227 to be used by popular CNNs. The database consists of 3 categories of images of chest X-rays, of which 219 are COVID-19 positive, 1345 are viral pneumonia and 1341 are normal chest X-rays. From the popular Kaggle database by Paul Moore, the normal along with the viral pneumonia images are sourced, ‘Chest X-Ray Images (Pneumonia)’. Whereas, the COVID-19 positive images are collected from various open sources including the Italian Society of Medical and Interventional Radiology (SIRM) COVID-19 Database, Novel Corona Virus 2019 (nCOVID-19) Dataset by Joseph Paul Cohen, Paul Morrison and Lan Dao, and 43 different publications as well. Figure 2 represents the sample images (X-ray) of the three classes obtained from chest X-ray database.

**Fig. 2.**
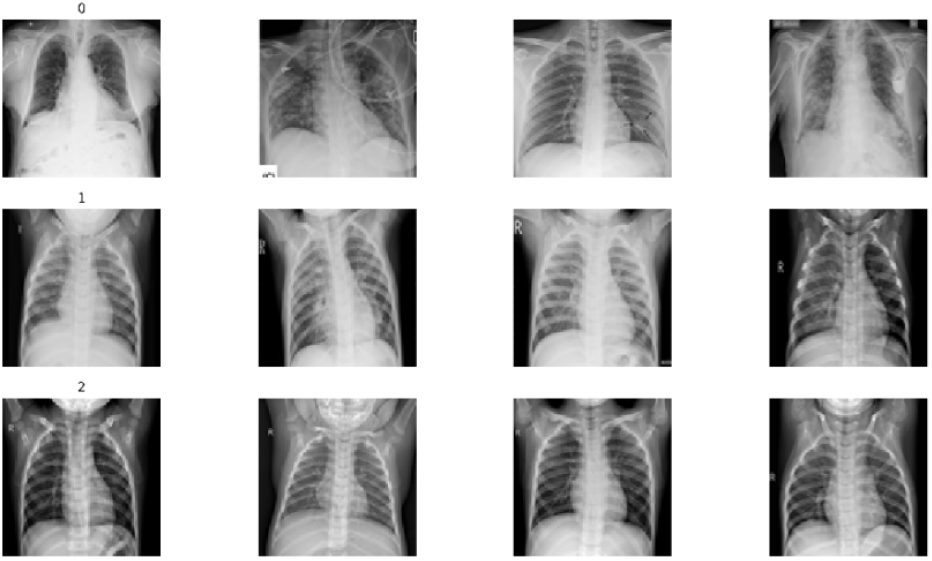
Sample X-ray images of three classes; 0: COVID, 1: Viral Pneumonia, 2: NORMAL from the chest X-ray database

### B. Data Pre-processing and Augmentation

Medical images are very often contaminated by noises due to increasing forms of intrusion including that of the procedure of imaging and data collection. As a consequence, a visual assessment of them could become more challenging. Several pre-processing techniques can be applied to enhance the information the image generates for the unaided eye or to use it as feedback for algorithms.

Firstly, the data samples are resized to 100×100 pixels and are converted to grayscale images. Each image is then merged into 3 channels resulting in an input shape of 100×100×3. Furthermore, the dataset is normalized using standardization or Z-normalization. Standardization, in machine learning algorithms, helps to stabilize the model as well as increases the speed of training. It is applied by the following equation,

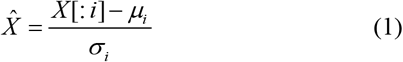

Additionally, the labels of the dataset (defining the category of the images) are one-hot encoded where each class (value of label) is converted into a binary feature. Augmentation of data is a method often applied in DL that helps to generate the number of samples required. It also enhances the effectiveness of the network for a small databases by optimizing it. Since our dataset is relatively small, we apply some image augmentation techniques for artificially increasing the size of our training data. To increase variation within our small database, we have applied image augmentations by gripping Keras *ImageDataGenerator* during training such as randomly rotating the images by 10°, randomly zooming images by a range of 0.9 to 1.1, as well as translating the images with respect to height and width proportional to a range of −0.1 to 0.1.

### C. Database Splitting

80:10:10 is the data splitting ratio used for the training, validation, and test sets of the dataset of 2905 images. The value counts of the respective datasets are as follows,

- Training Dataset: 2325 total images of which 175, 1077 and 1073 are the number of images (chest X-ray) of COVID-19 positive, viral pneumonia and normal respectively.
- Validation Dataset: 291 total images of which 22 are of COVID-19 positive, 135 are of viral pneumonia and 134 are of normal chest X-ray images.
- Testing Dataset: 291 total images of which 22 are of COVID-19 positive, 135 are of viral pneumonia and 134 are of normal chest X-ray images.

Table I depicts the distribution of images into training, validation, and test sets after splitting.

**TABLE I.**
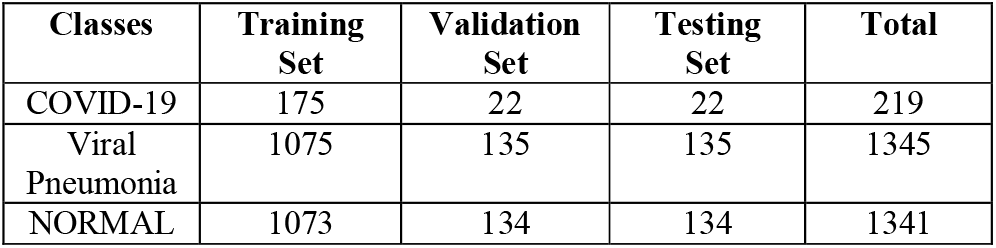
Distribution of chest X-ray Images After Splitting

| Classes | Training Set | Validation Set | Testing Set | Total |
| --- | --- | --- | --- | --- |
| COVID-19 | 175 | 22 | 22 | 219 |
| Viral Pneumonia | 1075 | 135 | 135 | 1345 |
| NORMAL | 1073 | 134 | 134 | 1341 |

### D. ResNet-50 and Transfer Learning

ResNet [11] has taken the turn of being the revolutionary deep neural network (DNN) model for computer vision tasks. Its breakthrough came in 2015 when it won the ImageNet [12] competition in 2015. Needless to say, DNNs perform better than neural networks (NN) with lesser layers in most cases. However, training a hugely stacked NN has been infamous for its vanishing gradient problem that causes performance deterioration in models. ResNet models, having up to 150+ layers, have solved this issue using identity shortcut connections – they’re connections that skip one or more layers. This provides a detour for gradients to pass through without diminishing. In this study, we have used the 50 layers ResNet-50 as the base architecture of the model and have fine-tuned it for our classification problem. The structure of the ResNet-50 used for the classification of chest X-ray is described in Figure 3.

**Fig. 3.**
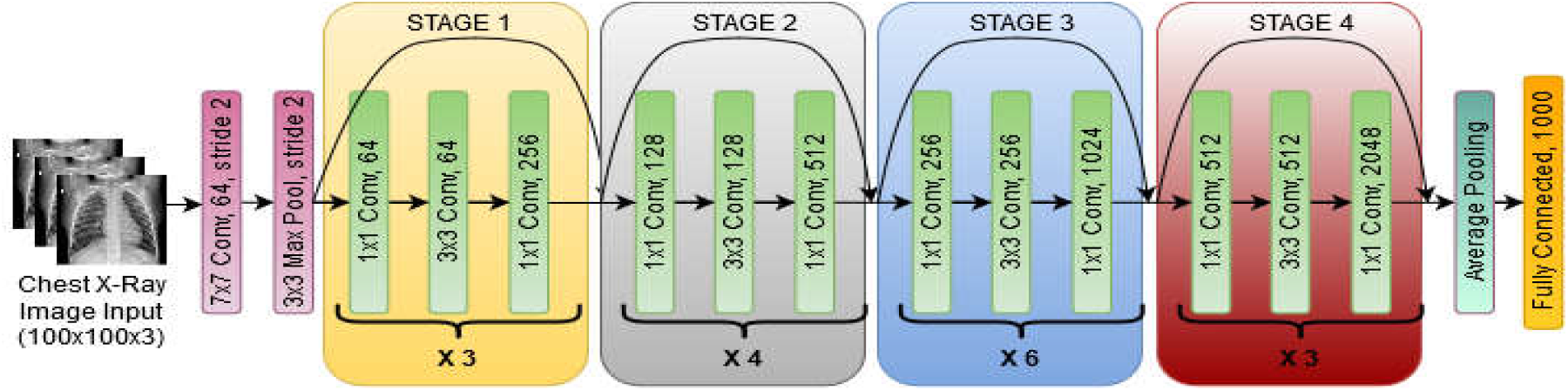
Structure of ResNet-50 for the classification of COVID-19 from chest X-ray images

Similar to how we use the knowledge of alphabets and learn to read words, models can also be created using the knowledge of neural networks that have been trained previously on similar data domain. They are useful because they have already learned features from a large database.

Since it doesn’t require us to build a deep neural network of such extensiveness from scratch, it can save us time and computational power. ResNet-50, pre-trained on ImageNet allows us to use its knowledge on a smaller database like ours. It already has learned patterns from many images [13]. Since the last layers detect task-specific patterns, we retrain it to be specific for our task. This concept is known as transfer learning [14].

### E. Proposed Transfer Learning with ResNet-50

The architecture of the proposed model for our task involves a ResNet-50 model followed by 4 additional task-specific layers. The weights of it pre-trained on ImageNet are loaded. The input size of the ResNet-50 is 100×100×3 and it uses average pooling. Figure 4 demonstrates the fine-tuned transfer learning on the ResNet-50 architecture.

**Fig. 4.**
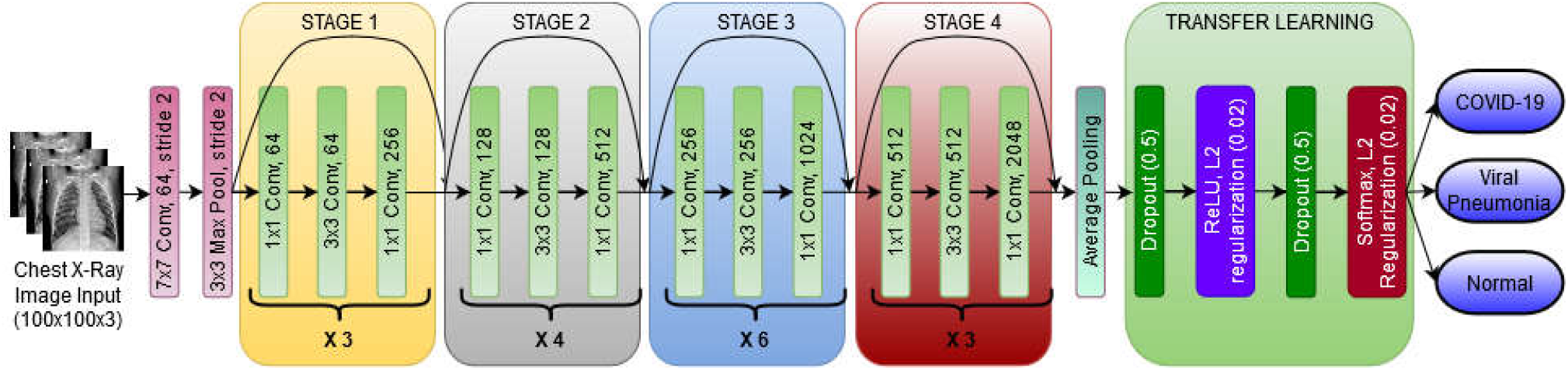
Proposed deep transfer learning on the ResNet-50 for the classification of COVID-19 from chest X-ray images

In this particular work, the DCNN model was fine-tuned and transfer learning applied for classifying images (chest X-ray) to detect COVID-19. The additional layers are replacing the fully connected layer of the ResNet-50. They are a dropout layer with a dropout probability of 50% - this drops 50% of the parameters randomly and reduces overfitting. It is followed by a Rectified Linear Unit (ReLU) activation layer with L2 regularization/ridge regression. The value of the regularization parameter is 0.02. Regularization is needed to prevent overfitting of the model, and it is done so by adding a penalty with the cost function. Next is another dropout layer with 50% dropout probability. And finally is a softmax activation layer with L2 regularization (regularization parameter = 0.02) and an output size of 3 for the 3 classes of COVID-19, viral pneumonia and normal.

## III. Experimental Results and discussion

### A. Experimental Setup

The chest X-ray dataset consists of X-rays of three types: COVID-19, Viral Pneumonia, and Normal. In our implemented fine-tuned DCNN using transfer learning, ResNet-50, 80% data was used for training, 10% of the data for validation, and the leftover 10% was for the testing. To train the neural network, Adam optimizer is employed as the optimization algorithm with the initial learning rate of 0.001 and categorical cross-entropy as the loss function for our multi-class classification problem. Moreover, we have used the Snapshot Ensemble approach during the training of our model. This approach allows us to control the learning rate in a way that helps us travel through all the local minima during the gradient descent algorithm and thus forming multiple models in one neural network. The learning rate starts at a fixed maximum value (of 0.001 in our case), and drops quickly near local minima, and subsequently jumps to the fixed maximum value again. We have selected to have an ensemble of 3 models in this task where each model is comprised of 20 epochs, i.e. we assist the model to converge to a local minimum within 20 epochs. Throughout our experiment, we used a workstation with the LINUX server (Ubuntu 16.04), NVIDIA GeForce GTX 2080ti GPU, 16GB RAM. The DCNN model was executed by the use of python in Keras package, running on TensorFlow backend on Intel Xeon E3, core i5-2.4GHz processor.

### B. Overall Performance Analysis

We assessed the performance of our deep transfer learning model for validation and testing dataset considering the following evaluation metrics: accuracy (ACC), precision (PPR), sensitivity or recall (SN), specificity (SP), and F1-score. The following equation measures the performance metrics,

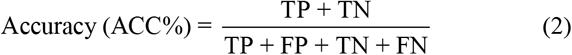

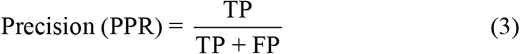

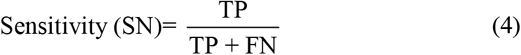

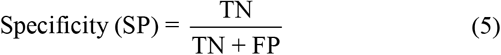

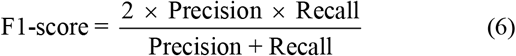

The model’s classification performance can be observed by the confusion matrices [15] provided in Figure 5.

**Fig. 5.**
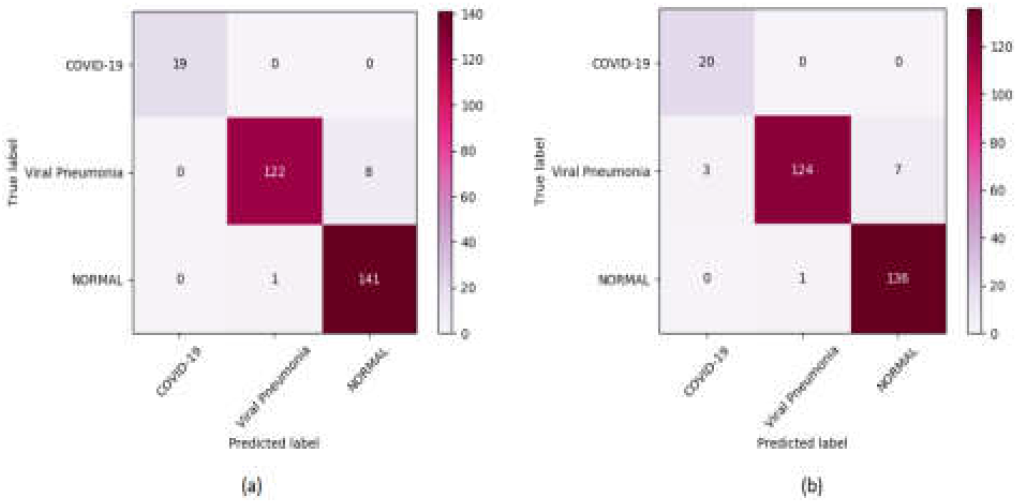
Confusion matrix of the proposed model: (a) test set (b) validation set

It is evident that the model is demonstrating a significantly good performance in case of detecting COVID-19 X-rays with correctly predicted fraction of 1 in both validation and test datasets. Bar plot in Figure 6 shows the correctly predicted fraction values for other cases as well.

**Fig. 6.**
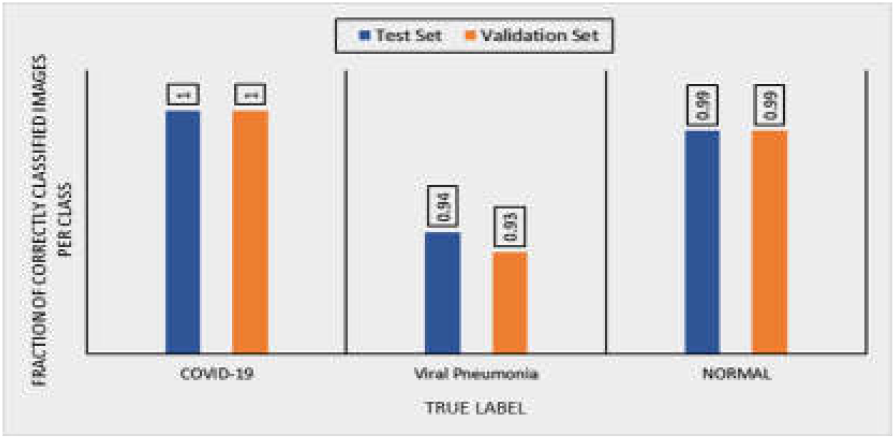
Bar plot demonstrating number of correctly predicted images in each class

As the dataset is imbalanced, rather than solely relying on the classification accuracy as a model performance evaluating metric, we considered precision, sensitivity, F1-score and specificity evaluation to justify the preeminence of the implemented model. The model performance is depicted in Table II.

**TABLE II.**
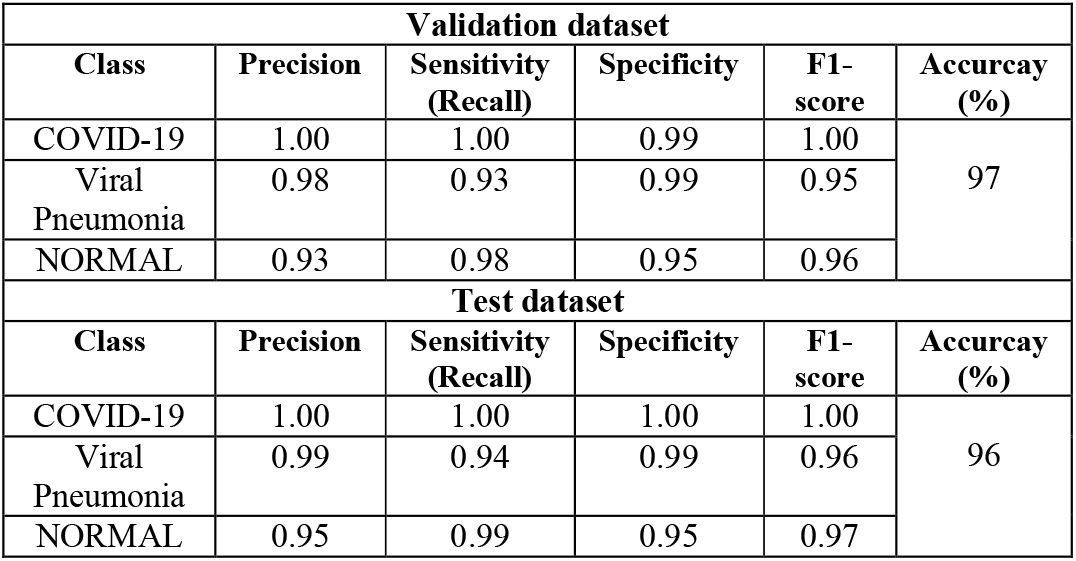
Performance Results of the COVID-19 Classification Obtained from Deep Transfer Learning Model

These performance evaluation parameters demonstrate that the model can detect COVID-19 cases with precision = 1.00, sensitivity (recall) = 1.00, specificity = 1.00, and F1-score = 1.00 scores on the testing dataset. Figure 7 and 8 illustrates the precision-recall relationship in individual cases on test and validation dataset.

**Fig. 7.**
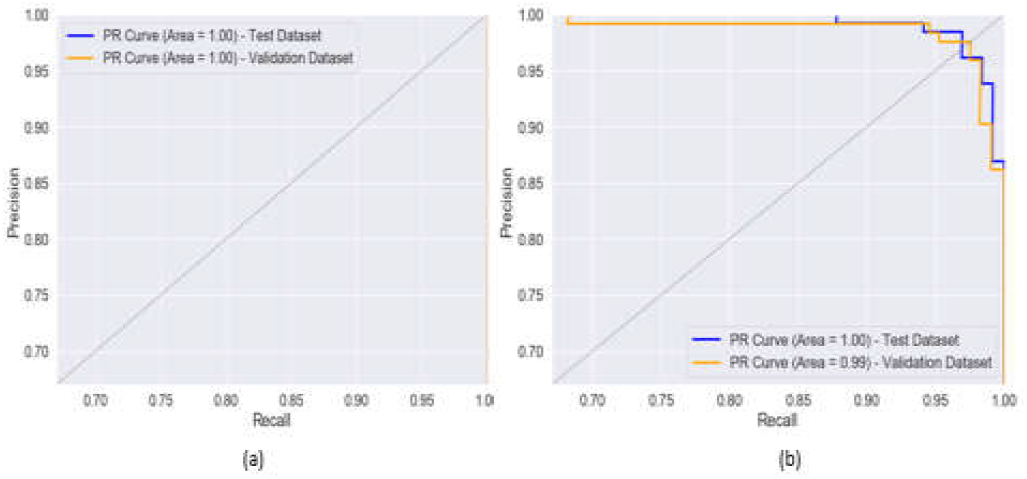
Precision-Recall curves for the class: (a) COVID-19 (b) Viral Pneumonia

**Fig. 8.**
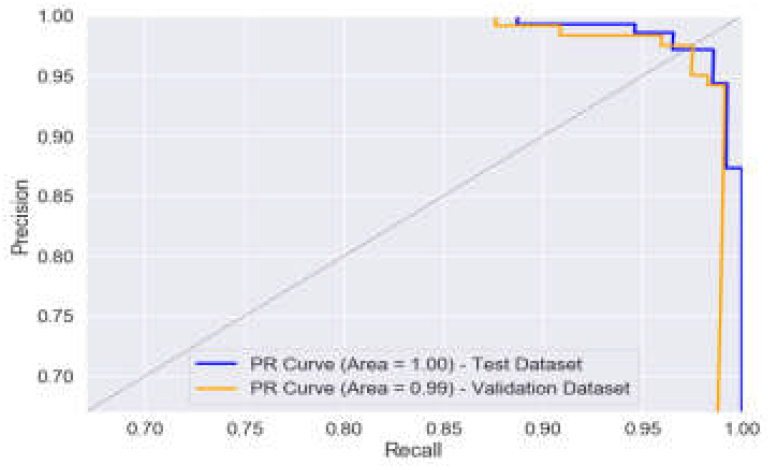
Precision-Recall curve for the class NORMAL

The ROC curves representing the TPR and FPR on test and validation dataset for the classes COVID-19, Viral Pneumonia and Normal X-ray, provided in Figure 9 and 10. Identification of COVID-19 from the X-ray images (chest) of viral pneumonia and healthy cases can be done by the model with reasonable accuracy (AUC=100%) as understood from the ROC curves. Therefore, the deep transfer learning model presented in this paper can be a reliable method for faster and accurate COVID-19 affected case detection. However, the model’s validation loss is 0.11 and the testing loss is 0.14. The training and validation loss is presented in Figure 11.

**Fig. 9.**
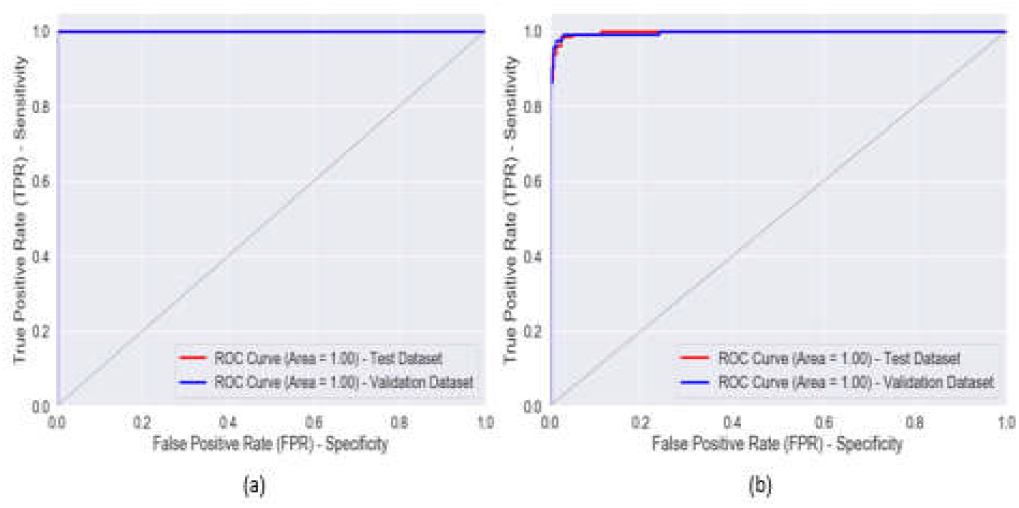
ROC curves representing the class: (a) COVID-19 (b) Viral Pneumonia

**Fig. 10.**
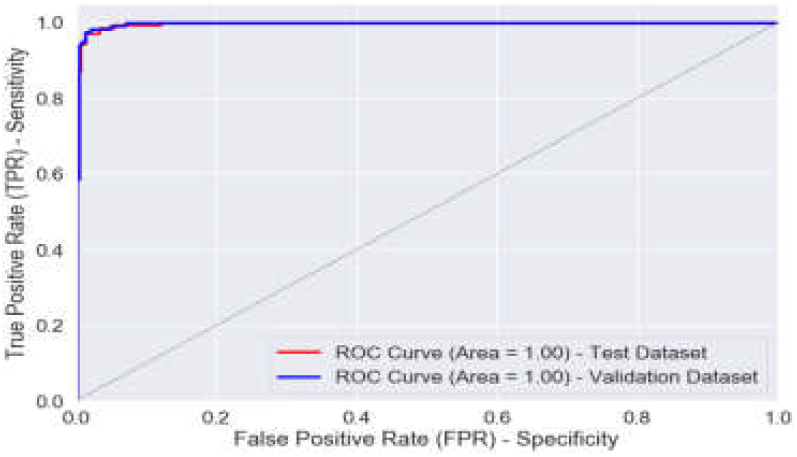
ROC curves representing the class NORMAL

**Fig. 11.**
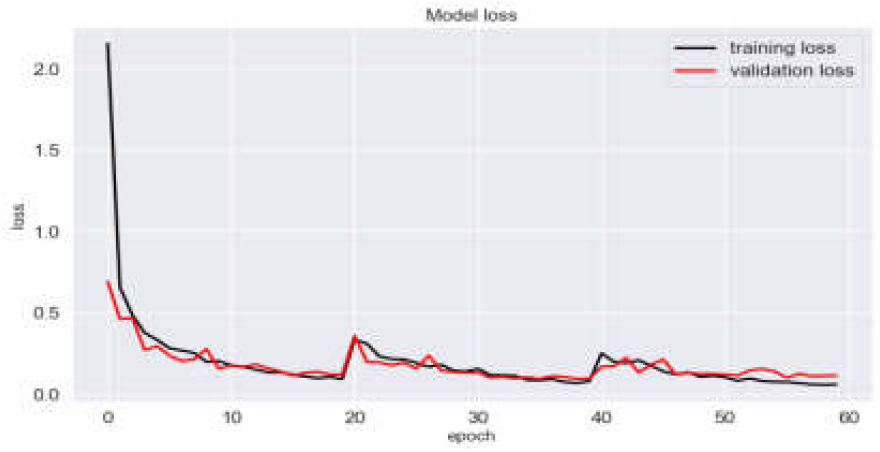
Loss between training and validation by ResNet-50 model

To evaluate our classification, we compared our proposed COVID-19 classifier with state-of-the-art approaches. Table III provides a comparison of our proposed model’s performance with the previously existing model. Our proposed model clearly outperforms some of the conventional methods in detecting COVID-19 by achieving 96.9% of average classification accuracy.

**TABLE III.**
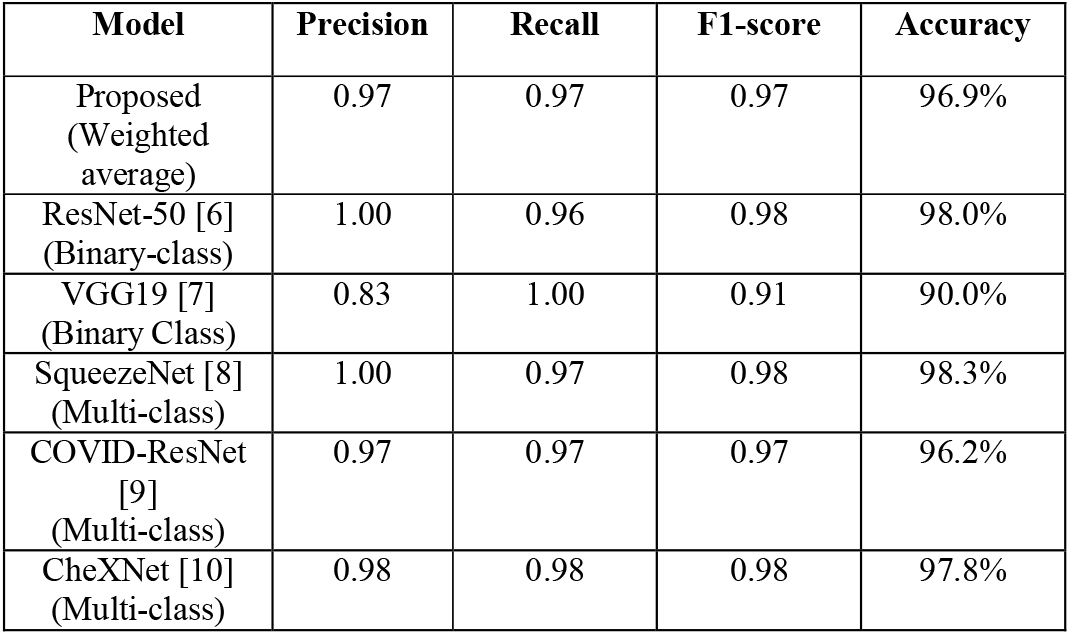
Performance Comparison of the Proposed COVID-19 Model with the Existing Deep Learning

## IV. Conclusion

In this study, a deep transfer learning model for detecting COVID-19 automatically from images (chest X-ray) along with two other classes (Viral Pneumonia and normal chest X-ray images). The model was trained using a pre-trained network ResNet-50. Later, transfer learning is applied to the pre-trained network for faster and efficient training which improved the performance of the model. The experimental analysis confirmed that the emerging deep neural network effectively performed in detecting COVID-19 from X-ray images as opposed to existing state-of-the-art methods. Several data augmentation and preprocessing have been performed in order to increase the size of the training dataset. Performance results demonstrate that the model achieved the validation and testing accuracy of 97% and 96% respectively. Moreover, the model exhibits exceptionally well in classifying the COVID-19 cases in the test dataset with a Precision of 1.00, Sensitivity 1.00, Specificity 1.00, and F1-score of 1.00. In conclusion, we believe that this transfer learning model used in the paper, as well as other presents in the literature, will help doctors to make decisions to detect COVID-19 at its rudimentary stage as the images (chest X-ray) pose an alternate option to the PCR method. In the future, we plan to extend our research by measuring the performance of the different algorithms using a large number of datasets.

## Data Availability

Not required

